# Early transmission dynamics of COVID-19 in a southern hemisphere setting: Lima-Peru: February 29^th^–March 30^th^, 2020

**DOI:** 10.1101/2020.04.30.20077594

**Authors:** César V. Munayco, Amna Tariq, Richard Rothenberg, Gabriela G Soto-Cabezas, Mary F. Reyes, Andree Valle, Leonardo Rojas-Mezarina, César Cabezas, Manuel Loayza, Peru COVID-19 working group, Gerardo Chowell

**Author notes:** Joint first authors (César V. Munayco, Amna Tariq). Corresponding author Amna Tariq, Department of Population Health Sciences, Georgia State University School of Public Health, Atlanta GA, 30303, Contact number.

## Abstract

The COVID-19 pandemic that emerged in Wuhan China has generated substantial morbidity and mortality impact around the world during the last four months. The daily trend in reported cases has been rapidly rising in Latin America since March 2020 with the great majority of the cases reported in Brazil followed by Peru as of April 15^th^, 2020. Although Peru implemented a range of social distancing measures soon after the confirmation of its first case on March 6^th^, 2020, the daily number of new COVID-19 cases continues to accumulate in this country. We assessed the early COVID-19 transmission dynamics and the effect of social distancing interventions in Lima, Peru.

We estimated the reproduction number, R, during the early transmission phase in Lima from the daily series of imported and autochthonous cases by the date of symptoms onset as of March 30^th^, 2020. We also assessed the effect of social distancing interventions in Lima by generating short-term forecasts grounded on the early transmission dynamics before interventions were put in place.

Prior to the implementation of the social distancing measures in Lima, the local incidence curve by the date of symptoms onset displays near exponential growth dynamics with the mean scaling of growth parameter, p, estimated at 0.9 (95%CI: 0.9,1.0) and the reproduction number at 2.3 (95% CI: 2.0, 2.5). Our analysis indicates that school closures and other social distancing interventions have helped slow down the spread of the novel coronavirus, with the nearly exponential growth trend shifting to an approximately linear growth trend soon after the broad scale social distancing interventions were put in place by the government.

While the interventions appear to have slowed the transmission rate in Lima, the number of new COVID-19 cases continue to accumulate, highlighting the need to strengthen social distancing and active case finding efforts to mitigate disease transmission in the region.

Peru COVID-19 working group

| Nombre | Area |
| --- | --- |
| Dante Castro Garro | Investigación Epidemiológica |
| Kevin Martel Vasquez | Investigación Epidemiológica |
| Edith Solis Castro | PREEC |
| Isabel Sandoval Ordinola | PREEC |
| Javier Masias Mimbela | PREEC |
| Kely Meza Cornejo | PREEC |
| Fabiola Caruajulca Quijano | PREEC |
| Lenin La Torre Rosillo | PREEC |
| Luis Ordoñez Ibarguen | PREEC |
| Mario Vasquez Dominguez | PREEC |
| Rommell Veintimilla Gonzalez Seminario | PREEC |
| Martha Calderón Silva | PREEC |
| Mayra Saavedra Dreyfus | PREEC |
| Miguel Luna Pineda | PREEC |
| Milagros Durand | PREEC |
| Noemí Janampa | PREEC |
| Jesus Chuquihuaccha | PREEC |
| Susan Mateo Lizarbe | PREEC |
| David Enriquez Cusi | PREEC |
| Idania Mamani Pilco | PREEC |
| Alex Jaramillo | PREEC |
| Karina Vargas | PREEC |
| Oswaldo Cabanillas | Alerta-Respuesta |
| Juan Arrasco | Alerta-Respuesta |
| María Vargas | Vigilancia en Salud Pública |
| Willy Ramos | Inteligencia sanitaria |

\* Corresponding author

## 1. Introduction

The Coronavirus disease 2019 (COVID-19) pandemic that emerged in the city of Wuhan in China in December 2019 has invaded nearly every nation of the world, becoming the most important public health emergency of the last century after the 1918–1920 influenza pandemic (WHO, 2020). In particular, the novel Severe Acute Respiratory Syndrome Coronavirus 2 (SARS-CoV-2) has an ability to exert substantial severe disease and mortality burden especially affecting individuals older than 60 years and those with prior health conditions including hypertension, cardiovascular disease, obesity and diabetes (Adler, 2020; Team, 2020). As of April 15^th^, 2020, the trajectory of the pandemic varies significantly around the world ranging from relatively well contained outbreaks in Thailand, Taiwan and Hong Kong to explosive epidemics characterized by initial exponential growth periods in a few hotspots located in various countries around the world including the United States, Italy, Spain, UK, France, and Iran (Ebbs, 2020; Griffiths, 2020; Minder, 2020).

By April 15^th^, 2020, SARS-CoV-2 is generating local transmission in over 200 countries and over 2.2 million cases and 150 thousand deaths have been reported globally (WHO, 2020). The COVID-19 pandemic was confirmed to have reached Latin America in February 2020 with a gradual expansion in the region until March 2020 when the COVID-19 incidence curve started to grow more rapidly. The US, the country with the highest number of reported COVID-19 cases in the world, has recorded 637,196 COVID-19 cases by April 15^th^, 2020. In Latin America, Brazil has reported 28320 cases, the highest number of cases in the region followed by Peru with a total of 11475 cases (Worldometer, 2020).

Peru, a country located in western South America, reported its first imported case of COVID-19 in Lima, on March 6^th^, 2020, a Peruvian with recent travel history to France, Spain and Czech Republic (Aquino & Garrison, 2020). By April 15^th^, 2020, a total of 11475 cases including 254 deaths have been reported by the Peruvian government. Lima, the capital of Peru has recorded 8412 cases, the highest number of cases within Peru (MOH, 2020). To respond to the growing number of COVID-19 cases in the country, the government shuttered schools on March 11^th^, 2020. The next day, the government banned gatherings of more than 300 people and suspended all international flights from Europe and Asia. On March 16^th^, 2020, the government declared a national emergency and closed country borders (Explorer, 2020). Subsequently, on March 17^th^, 2020 the president of Peru announced the beginning of community transmission of SARS-CoV-2 in the country, and ordered a curfew in the region on March 18^th^, 2020 to avoid night time socializing to prevent disease transmission (Explorer, 2020; Writing, 2020).

In order to combat the spread of the COVID-19 epidemic in Lima, the capital and largest city of Peru, estimates of the transmission potential of COVID-19 can guide the intensity of interventions including the reproduction number, R, during the early transmission phase (Nishiura & Chowell, 2009, 2014). Moreover, using the epidemiological data and mathematical modeling, it is possible to gauge the impact of control interventions including school closures and a national emergency declaration in Lima by assessing short-term forecasts grounded on the trajectory of the epidemic prior to the implementation of control interventions (Funk, Camacho, Kucharski, Eggo, & Edmunds, 2018; Shanafelt, Jones, Lima, Perrings, & Chowell, 2018).

## 2. Methods

### 2.1. Data

We analyzed the daily number of COVID-19 confirmed cases by date of symptoms onset in Lima, Peru by March 30^th^, 2020. Individual-level case details including whether the case was locally acquired or imported were also made available from the Centro Nacional de Epidemiología Prevención y control de Enfermedades and the National Institute of Health of the Ministry of Health, Peru (Group, 2020). We also examined the daily testing rate and the positivity rate from the daily number of positive and negative PCR test results by the date of reporting until March 30^th^, 2020.

### 2.2. Early growth model

We generate short-term forecasts in real time using the generalized growth model (GGM) that relies on two parameters and characterizes the early ascending phase of the epidemic allowing to capture a range of epidemic growth profiles including sub-exponential (polynomial) and exponential growth. GGM characterizes epidemic growth by estimating two parameters (i) the intrinsic growth rate, r and (ii) a dimensionless “deacceleration of growth” or “scaling of growth” parameter, p. The latter parameter modulates the epidemic growth patterns including the sub-exponential growth (p<1) and exponential growth dynamics (p=1). The GGM model is given by the following differential equation:

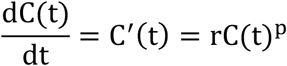

Where C′(t) describes case incidence over time t. The cumulative number of cases at time *t* is given by *C*(*t*) while *r* is a positive parameter denoting the growth rate (1/time) and *p*∈[0,1] is a “deceleration of growth” parameter (Chowell, 2017; Viboud, Simonsen, & Chowell, 2016).

### 2.3. Short term forecast to assess interventions

We calibrate the GGM model to the daily case incidence by the date of symptoms onset for Lima. We analyzed the time series data of confirmed cases by onset dates for Lima from February 29^th^, 2020 to March 30^th^, 2020. Our model was calibrated using case series from February 29^th^–March 15^th^, 2020, prior to the implementation of national emergency in Lima.

The best fit model solution is estimated by using a non-linear least square fitting approach (full details provided in (Chowell, 2017)). This process searches for the set of model parameters that minimizes the sum of squared differences between the observed data y_ti_ = y_t2_, …. y_tn_ and the corresponding model solution given by f(t_i_, Θ): where Θ = (r, p) correspond to estimated set of parameters of the GGM model. Thus, the objective function for the best fit solution of f(t_i_, Θ) is given by

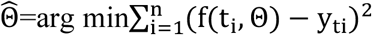

The initial condition is fixed to the first observation in the data. Next, we derive uncertainty around the best fit model solution as well as the confidence intervals of the parameters utilizing a parametric bootstrapping approach assuming a Poisson error structure as described in ref (Chowell, 2017).

### 2.4. Reproduction number from case incidence using the generalized-growth model

Generally, the reproduction number, R, quantifies the average number of secondary cases per case during the early ascending phase of an outbreak before the implementation of interventions or behavior changes (Anderson & May, 1991; Chowell et al., 2015; Yan & Chowell, 2019). Estimates of the effective R indicate if the disease transmission sustains (R>1) or the disease trend is declining (R<1). Therefore, it is necessary to maintain R<1 to contain an outbreak. Here, we estimate the reproduction number by characterizing the early growth phase (16 day) of local cases using the generalized-growth model (Viboud et al., 2016) and modeling the generation interval of SARS-CoV-2 assuming a gamma distribution with a mean of 4.41 days and a standard deviation of 3.17 days (Nishiura, Linton, & Akhmetzhanov, 2020; You et al., 2020). We simulate the progression of local incident cases by onset dates using the calibrated GGM model and account for the daily series of imported cases into a renewal equation given as (Nishiura & Chowell, 2009, 2014; Paine et al., 2010):

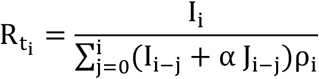

We denote the local incidence at calendar time t_i_ by I_i_, which is characterized using the generalized-growth model, the imported cases at calendar time t_i_ by J_i_, and the discretized probability distribution of the generation interval by ρ_i_. In this equation the numerator represents the total new cases I_i_, and the denominator represents the total number of cases that contribute to the new cases I_i_ at time t_i_. The relative contribution of imported cases to the secondary disease transmission is represented by the parameter 0≤ α ≤1. We perform a sensitivity analyses by setting α = 0.15 and α = 1.0 to assess the relative contribution of imported cases to the secondary disease transmission (Nishiura & Roberts, 2010). This is followed by the derivation of the uncertainty bounds around the curve of R directly from the uncertainty associated with the parameter estimates (r, p). We estimate R for 300 simulated curves assuming a Poisson error structure (Chowell, 2017). This method to derive early estimates of the reproduction number, R, has been employed in several prior studies as in refs (Chowell, 2017; Tariq et al., 2020).

## 3. Results

### 3.1. COVID-19 testing and positivity rates

Figure 1 shows the daily number of positive and negative laboratory test results and the positivity rate during the reporting period, March 4^th^–March 30^th^, 2020. The total number of PCR tests performed for this time period were 11518 (1127 positive results and 10307 negative results). The average daily number of PCR tests performed in Lima was estimated at ~188 between March 4^th^–March 15^th^, 2020 whereas the number of tests performed between March 16^th^–March 30^th^, 2020 increased to ~617 tests per day, an increase of 228 % in the testing rates, perhaps reflecting an increase number of suspected cases with respiratory symptoms. The positivity rate (percentage of positive cases among the positive and negative cases) has ranged from 0.6-23.9 % between March 4^th^–March 30^th^, 2020.

**Figure 1:**
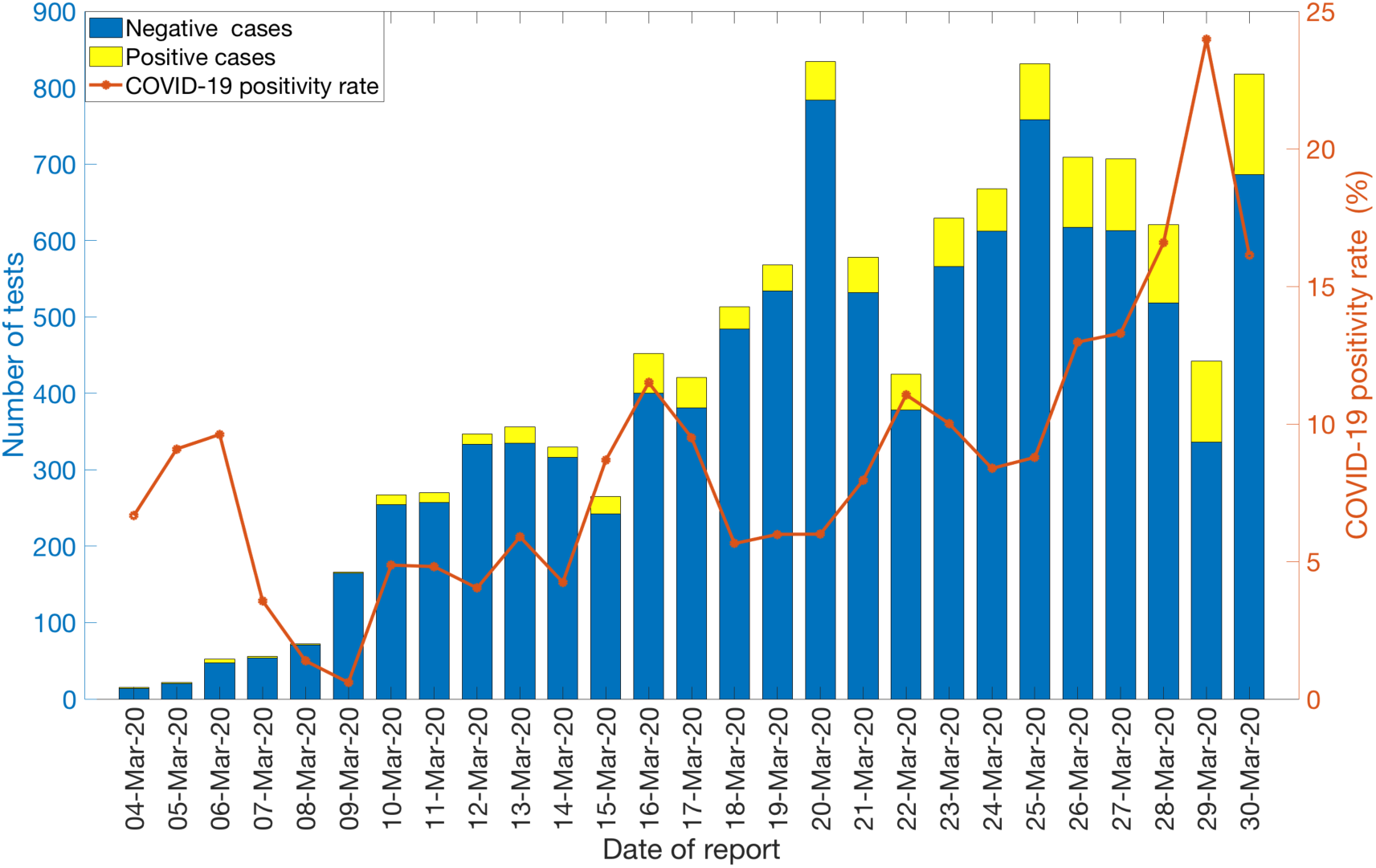
Laboratory results of COVID-19 tests in Lima as of March 30^th^, 2020. Blue color represents the negative test results and the yellow color represents the positive test results. The orange solid line denotes the COVID-19 positivity rate in Lima.

### 3.2. Local and imported incidence trends

The COVID-19 epidemic curve by the date of symptoms onset, stratified by the local and imported incidence case counts is shown in Figure 2. On average ~6 imported cases and ~162 local cases have been reported daily between March 16^th^–March 30^th^, 2020 in Lima. A total of 2783 autochthonous cases and 151 imported cases have been reported in Lima as of March 30^th^, 2020.

**Figure 2:**
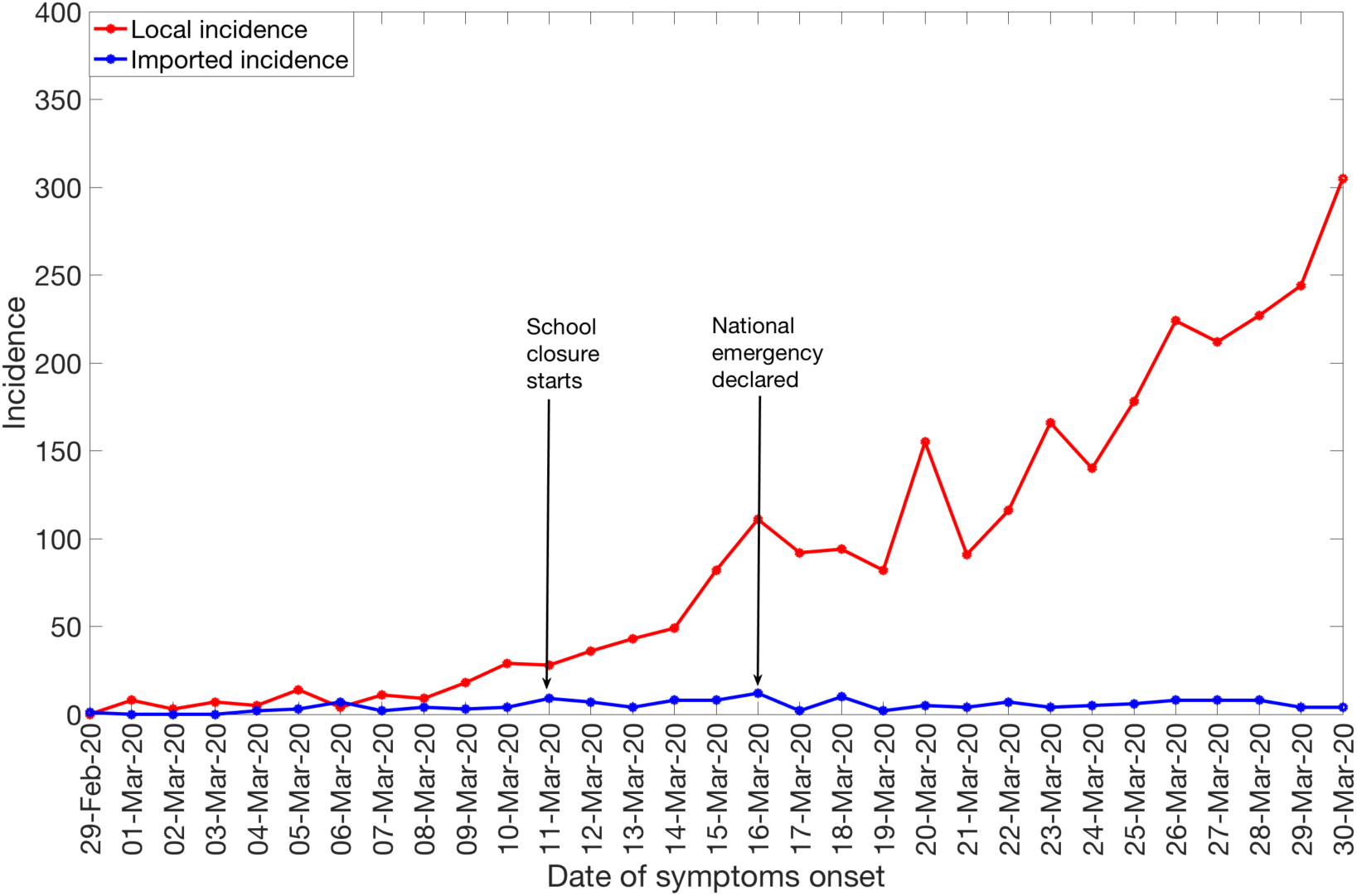
Daily numbers of new local and imported confirmed COVID-19 cases in Lima by date of symptoms onset as of March 30^th^, 2020.

### 3.3. Reproduction number, R

We estimated the reproduction number from the epidemic’s early growth phase comprising the first 16 epidemic days prior to the implementation of social distancing interventions which includes the national emergency declaration on March 16^th^ 2020. The local incidence curve by the date of symptoms onset displays near exponential growth dynamics with the scaling of growth parameter, p, estimated at 0.9 (95% CI: 0.9, 1.0) and the intrinsic growth rate, r, estimated at 0.3 (95% CI: 0.3, 0.5). The estimate of the reproduction number was estimated at 2.3 (95% CI: 2.0, 2.5) when α = 0.15 (Figure 3). When α = 1.0, the reproduction number slightly decreases to 2.0 (95% CI: 1.7, 2.3) (Table 1).

**Table 1:** Mean estimates and the corresponding 95% confidence intervals for the reproduction number in Lima, growth rate and the scaling of growth parameter during the early growth phase as of March 15^th^, 2020

| Parameter | Estimated values at<br>$\alpha = 1.0$ | Estimated values at $\alpha =$<br>**0.15** |
| --- | --- | --- |
| **Reproduction number** | 2.0 (95% CI: 1.7,2.3) | 2.3 (95%CI: 2.0,2.5) |
| **Growth rate, r** | 0.3 (95%CI: 0.3,0.5) |  |
| **Scaling of growth parameter, p** | 0.9 (95%CI: 0.9,1.0) |  |

**Figure 3:**
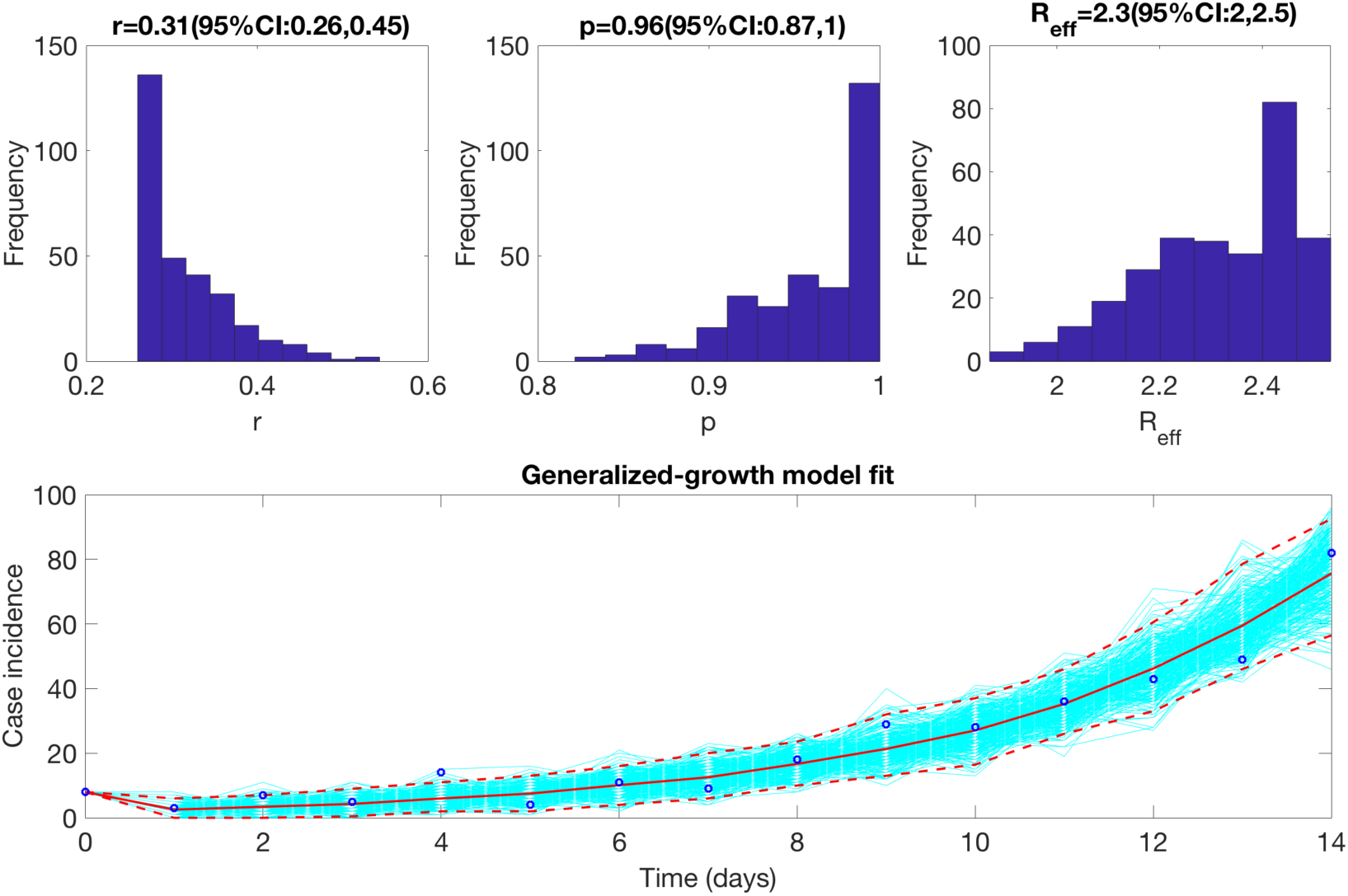
The reproduction number derived from the early growth phase in the number of COVID-19 cases in Lima after adjusting for imported cases with *α* = 0.15 using the GGM model as described in the text. The reproduction number based on the incidence curve by March 15^th^, 2020 was estimated at 2.3 (95% CI: 2.0, 2.5).

### 3.4. Assessing the impact of social distancing interventions

In order to assess the impact of social distancing interventions in Lima, including school closures on March 11^th^, 2020 and the declaration of national emergency on March 16^th^, 2020, we generate a 20-day ahead forecast for Lima based on the daily incidence curve up until the declaration of the national emergency in Lima. The 16-day calibration period of the model yields an estimated growth rate, r, at 0.8 (95% CI: 0.6, 1.1) and a scaling of growth rate parameter, p, at 0.8 (95%CI: 0.7,0.9). The 20-day ahead forecast suggests that the effect of the school closure and the national emergency declaration slowed down the spread of the virus as shown in Figure 4. Indeed, the scaling of growth parameter declined to 0.53 (95% CI: 0.48, 0.58), consistent with an approximately linear incidence growth trend during the period affected by the intervention.

**Figure 4:**
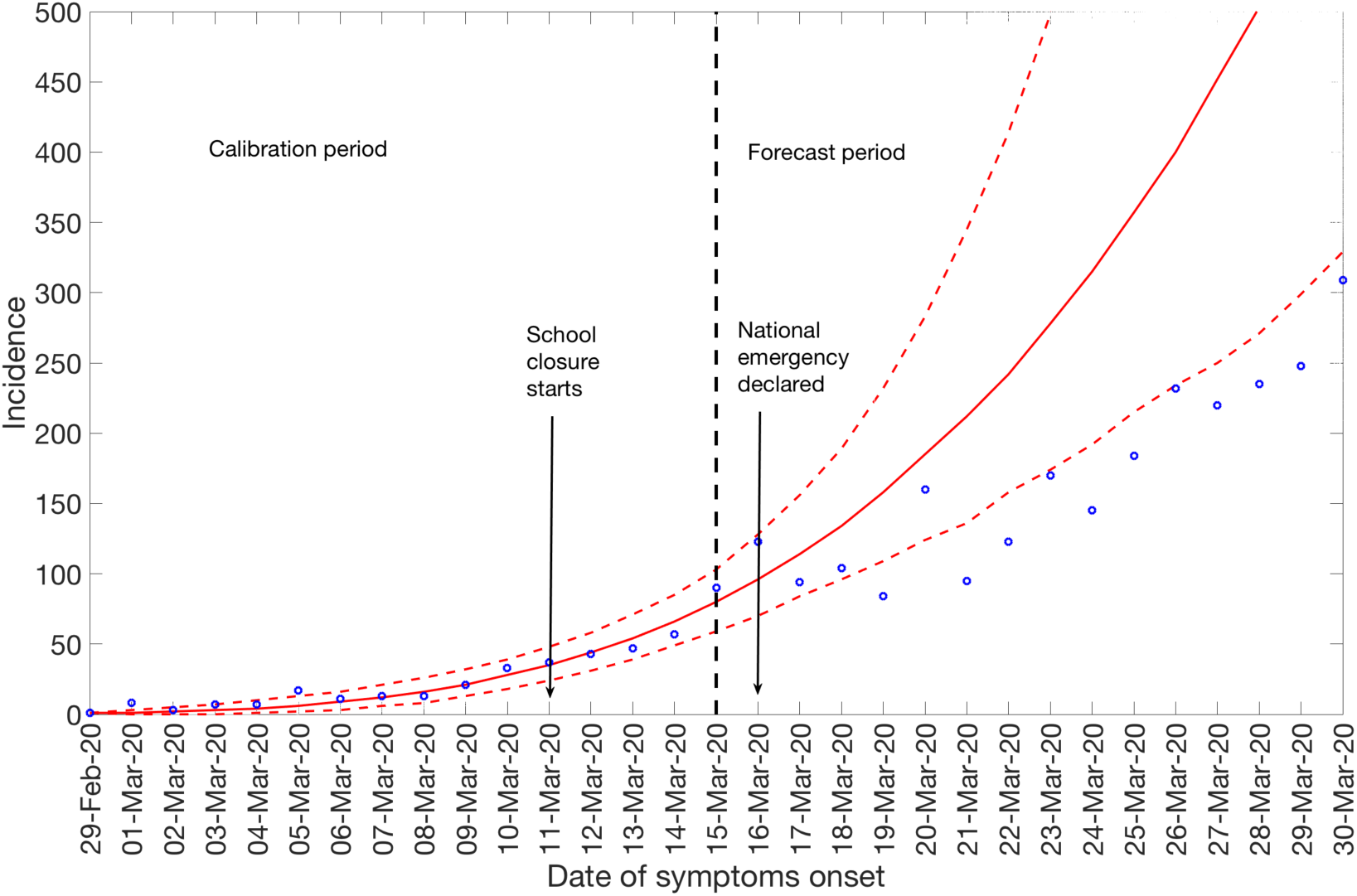
20-day ahead forecast of the COVID-19 epidemic in Lima by calibrating the GGM model until March 15^th^, 2020 (vertical dashed line). Blue circles correspond to the data points, the red solid line indicates the model’s mean fit and the red dashed lines represent the 95% prediction interval. The vertical black dashed line represents the time of the start of the forecast period. The forecast (March 16^th^- March 30^th^) suggests that social distancing interventions have slowed down the transmission rate.

## 4. Discussion

Our estimate of the transmission potential in Lima for the first 16 days of the epidemic indicates sustained local transmission in the region after accounting for multiple case importations with the estimate of reproduction number, R, at ~2.3 (95% CI: 2.0, 2.5) which is comparable to estimates of the reproduction number for China, Korea, and Iran that lie in the range of 1.5-7.1 (Hwang, Park, Kim, Jung, & Kim, 2020; Mizumoto, Kagaya, & Chowell, 2020; Muniz-Rodriguez et al., 2020; Read, Bridgen, Cummings, Ho, & Jewell, 2020; Shim, Tariq, Choi, Lee, & Chowell, 2020; Wu, Leung, & Leung, 2020). In contrast, a recent study on Singapore’s COVID-19 transmission reported a lower estimate of R at ~0.7, which has been explained as a result of the early implementation of sweeping social distancing interventions (Tariq et al., 2020).

The initial scaling of growth parameter in Lima indicates a nearly exponential growth pattern, consistent with the early spread of the COVID-19 epidemic in Iran and the exponential growth pattern of COVID-19 displayed by the Chinese province of Hubei (Muniz-Rodriguez et al., 2020; Roosa et al., 2020). In comparison sub-exponential growth patterns of COVID-19 have been observed in Singapore (p~0.7), Korea (p~0.76) and other Chinese provinces excluding Hubei (p~0.67) as described in recent studies (Roosa et al., 2020; Shim et al., 2020; Tariq et al., 2020).

Although Lima has been quick to take aggressive measures against COVID-19, Peru remains one of the hardest hit countries in Latin America (Tegel, 2020). Despite the closure of country borders on March 16^th^, 2020, the number of imported cases in Lima has increased with an average of ~6 imported cases reported between March 16^th^–March 30^th^, 2020 compared to an average of ~4 imported cases per day before March 16^th^, 2020. However, the 20-day ahead forecast of our GGM model calibrated to first 16 epidemic days suggest that the social distancing measures, including closure of schools and the declaration of national emergency are slowing down the virus spread in Lima. The scaling of growth parameter, p, was estimated at ~0.5 (95%CI: 0.5,0.6) after the implementation of social distancing measures, consistent with a linear incidence growth trend. However, the COVID-19 case incidence continues to accumulate despite the quarantine and lockdowns in the region highlighting the need to enhance social distancing measures to further contain the outbreak.

The average positivity rate of COVID-19 in Lima was ~8.6% between March 4^th^–March 30^th^, 2020. This positivity rate for Lima, Peru, corresponds to the positivity rates derived from Denmark, Germany and Canada (6-8%) (Meyer & C.Madrigal, 2020). In comparison countries like New Zealand, South Korea and Australia have tested widely and exhibit lower positivity rates (2%) whereas Italy and the US have shown much higher positivity rates (15-20%) for COVID-19 indicating suboptimal testing capacity in these countries (Meyer & C.Madrigal, 2020; Project, 2020). A recent study has shown that changes in testing rates over the course of the epidemic can mask the epidemic growth rate resulting in biased epidemic trends (Omori, Mizumoto, & Chowell, 2020). Moreover, there is a substantial fraction of asymptomatic COVID-19 cases, which could have underestimated the reproduction number derived from the daily incidence’s growth trend of symptomatic cases (Mizumoto, Kagaya, Zarebski, & Chowell, 2020; Wei et al., 2020). Our study underscores the need for active contact tracing efforts that targets symptomatic and asymptomatic cases, rapid isolation of infectious individuals, quarantined contacts and strict social distancing measures to curb the spread of the virus.

## 5. Conclusion

In this study we estimate the early transmission potential of SARS-CoV-2 in Lima, Peru. Our current findings point to sustained transmission of SARS-CoV-2 in the early phase of the outbreak, with our estimate of the mean reproduction number ~2.3. The COVID-19 epidemic in Lima followed an early exponential growth trend, which slowed down and turned into an almost linear growth trend (p~0.5), which appears to be tied to broad scale social distancing interventions put in place by the government. While the interventions appear to have slowed the transmission rate, the number of new COVID-19 cases continue to accumulate, highlighting the need to continue social distancing and active case finding efforts to mitigate disease transmission in the region.

## Data Availability

The data is not publicly available.

## List of abbreviations

COVID-19

SARS-CoV-2

PCR

## Author Contributions

A.T., C.M. and G.C. analyzed the data. A.T. and C.M. retrieved and managed data. A.T and G.C. wrote the first draft of the manuscript. A.T. and G.C worked on subsequent versions of the manuscript. All authors contributed to writing and interpretation of results. All authors read and approved the final manuscript.

## Ethical approval

Data has been made available and approved for analysis by the Centro Nacional de Epidemiología Prevención y control de Enfermedades (CDC Perú) and the National Institute of Health of the Ministry of Health, Peru.

